# COVID-Anosmia Checker: A rapid and low-cost alternative tool for mass screening of COVID-19

**DOI:** 10.1101/2020.10.28.20221200

**Authors:** Budhaditya Basu, Paul Ann Riya, Joby Issac, Surendran Parvathy, Biju Surendran Nair, Pradipta Tokdar, Devika Sanal Kumar, Pranav Ravi Kulkarni, Gowtham Hanumanram, Mohanan Jagadeesan, Prasanna Karthik Suthakaran, Lal Devayani Vasudevan Nair, Rosy Vennila, Rajendran Kannan, Balarama Kaimal, Gopa Kumar Anoop, Iype Joseph, Radhakrishnan Nair, Saji George, Jackson James, Madhavan Radhakrishna Pillai

## Abstract

**Background:** COVID-19 curve can be flattened by adopting mass screening protocols with aggressive testing and isolating infected populations. The current approach largely depends on RT-PCR/rapid antigen tests that require expert personnel resulting in higher costs and reduced testing frequency. Loss of smell is reported as a major symptom of COVID-19, however, a precise olfactory testing tool to identify COVID-19 patient is still lacking.

**Methods:** To quantitatively check for the loss of smell, we developed an odor strip, “*COVID-Anosmia checker”*, spotted with gradients of coffee and lemon grass oil. We validated its efficiency in healthy and COVID-19 positive subjects. A trial screening to identify SARS-CoV-2 infected persons was also carried out to check the sensitivity and specificity of our screening tool.

**Findings:** It was observed that COVID positive participants were hyposmic instead of being anosmic when they were subjected to smelling higher odor concentration. Our tool identified 97% of symptomatic and 94% of asymptomatic COVID-19 positive subjects after excluding most confounding factors like concurrent chronic sinusitis. Further, it was possible to reliably predict COVID-19 infection by calculating a loss of smell score with 100% specificity. We coupled this tool with a mobile application, which takes the input response from the user, and can readily categorize the user in the appropriate risk groups.

**Conclusion:** Loss of smell can be used as a reliable marker for screening for COVID-19. Our tool can be used for first-line screening to trace out COVID-19 infection effectively. It can be used in difficult to reach geographical locations.

## Introduction

The recent ongoing pandemic of COVID-19 caused by the Severe Acute Respiratory Syndrome Corona Virus 2 (SARS-CoV-2) has posed an enormous challenge to the human race^1,2^. It is important to note that the SARS-CoV-2 is more infectious than that of SARS-CoV and MERS-CoV since all infected individuals can potentially transmit the virus including those who are asymptomatic ^3–6^. That is why the overall number of deaths from COVID-19 outweighs that from SARS and MERS. Although at what stage and how frequently the asymptomatic individuals potentially transmit the disease remains to be elucidated. As on 24^th^ October 2020, the number of confirmed COVID-19 cases has risen to 42.4 million with more than 1.14 million deaths worldwide (https://coronavirus.jhu.edu/map.html). It is highly likely that the number of cases would be significantly greater than that reported because very mild or asymptomatic cases are excluded as they go untested. Several serological studies have shown that the number of infected cases is more than ten times higher than the confirmed cases ^7–9^. Thus, it raises the question of the efficiency of the broad screening methods adopted currently to prevent the spread of the disease. Given that the COVID-19 cases can surge as a second wave, it is high time to switch from the highly sensitive PCR based surveillance regime to a sufficiently inexpensive and easy to test regime. Perhaps a test that can be performed at home multiple times a week would better serve the purpose ^10,11^.

The common symptoms of COVID-19 are fever, cough, fatigue and less common symptoms are headache and diarrhea^12,13^. But recent reports suggest that the loss of smell is a prominent symptom of COVID-19 infection ^14–20^ which is consistent with the listed symptoms by the Center for Disease Control and Prevention (Atlanta, GA, USA) ^21^. The identification of the impairment of sense of smell is particularly important in a country like India and similarly for other low and middle-income countries where mass screening through RT-PCR or antigen test is resource-intensive. Therefore, the approach should be made such that the patients with recent onset of olfactory dysfunction (OD), with or without other symptoms of COVID-19, should undergo self-isolation and, when possible, take confirmatory SARS-CoV-2 testing.

However, self-testing of olfactory function is difficult to standardize across locations, cultures and scenarios by asking participants to test themselves with household items like flowers, soap etc.^17^. Clearly quantitative assessment of smell perception (e.g., hyposmia) is not feasible in the case of self-reporting. Moreover, subjective bias is an added disadvantage to this way of assessing smell perception dysfunction. Here, we developed an olfactory assessment tool that can reliably test the participants without having a subjective bias. This tool seeks to assess a combination of parameters that includes the odor threshold (minimum strength of an odor that can be perceived), odor discrimination (differentiation between different odors) and odor identification. Moreover, this tool is also capable of identifying a quantitative reduction in smell (anosmia/hyposmia) as well as qualitative changes in smell (e.g., distortions of smell termed as parosmia, or phantom sensation termed phantosmia). This low-cost strip-based anosmia-screening tool can be used for mass screening of COVID-19 and here, we have validated its potential to be used as a first-line screening tool to trace out COVID-19 infection effectively.

## Methods

### Study Design

Participation in this study was voluntary and participants received no incentives. Inclusion criteria were: Age 20-60 years, with an oral consent to participate. Both males and females were included in the study. Exclusion criteria were: Age below 20 years and more than 60 years, patients with chronic sinusitis. Chronic COVID −19 positive patients with oxygen/ventilator support in ICU were excluded. The study was approved by the Institutional Human Ethics Committee of Rajiv Gandhi Center for Biotechnology (RGCB/IHEC/250/2020/28) and Saveetha Medical College Hospital (002/08/2020/IEC/SMCH).

### Sample size estimation

Sample size estimation was carried out in order to determine the study population. It was estimated that more than 25 participants were needed to have a confidence level of 95% with 20% marginal error. Population proportion by default was chosen as 50%.

### Participants

Participants who completed all the mandatory queries (that addresses odor identification, RT-PCR/Antigen/Antibody test results) were included in the present study. Participants who did not complete all required fields and/or provided incomplete responses in smell perception were excluded from the sample.

Here, we categorized the participants into three diagnostic groups: 1) Control participants who had no respiratory illness and were confirmed to be antigen or antibody (both IgM and IgG) negative to SARS-CoV-2 (N= 35; 9 M, 26 F; age range 25-50). These participants had not contacted any COVID-19 positive patients and had no COVID-like symptoms reported at the time of testing. 2) Participants who were tested RT-PCR positive for SARS-CoV-2 RNA upon nasopharyngeal or throat swab that were performed according to the WHO recommendations (N=95: 80 M, 15 F; age range 20-60). This group included both symptomatic and asymptomatic patients admitted at Saveetha Medical College Hospital, Chennai, India. 3) Suspected COVID-19 cluster (N=28; 5M, 23F; age range 20-35) and fever clinic outpatients (N= 30; 24M, 6F; age range 20-50) whose COVID-19 RT-PCR status was unknown at the time of olfactory assessment but was followed by RT-PCR test.

### Fabrication of COVID-Anosmia checker

*COVID-Anosmia checker* consisted of Part-A and Part-B **(Fig. S1A&B)**. Part-A consisted of the test strip as depicted **(Fig. S1A)**. The test strip consists of six black printed regions where positions 1, 3, 4, 5 contain different concentrations of coffee oil (2.0µl, 1.0µl, 2.5µl, and 5.0µl respectively, meant for odor threshold/ quantitative smell assessment). Position-2 was intentionally kept blank (to identify odor discrimination/phantosmia and also acted as placebo), and Position-6 contained lemongrass oil (5.0µl, for odor discrimination/quantitative smell assessment). This combination is ideal for assessing the quantitative reduction and qualitative changes in smell. Coffee oil and lemongrass oil is used since these two odors are very common to households and consistent with the previous self-reporting based study ^22^. The concentration of gradient and the pattern of spotting were standardized after a series of tests on normal healthy individuals. 1μl of coffee oil was the minimum volume that could be detected by any healthy individual. The spotted strip (odorized/non-odorized) was then wrapped with two layers of lamination to trap and stabilize the odor. The test strip with a combination of odor remained unknown to the subject being tested. The test subject had to cut each position separately along the dotted lines as depicted, smell the cut ends immediately and report the smell and intensity before proceeding to the next position **(Fig. S1A**).

The scoring sheet (Part-B) provided along with each paper strip sought to gather a few details of the participants after getting oral consent for their participation in the study **(Fig. S1B**). Section-1 includes the personal information and travel history of the test subject. In Section-2 the test subjects had to score Yes/No and mention the perceived odor in the dedicated column after smelling the *COVID-Anosmia checker* strip. Section-3 covers the symptoms if any, experienced by the test subjects and the date of onset of that particular symptom. Further an android based mobile application was developed to replace Part-B for easy decision making. Currently, the application is downloadable (https://neologix.ae/covid-anosmia-checker-app.html) to any android based smartphone and can be combined with test strips (Part-A) for self-testing or community screening.

### Smell Assessment criteria

Participant’s responses while identifying coffee and lemongrass oil varied depending upon their prior experience with that odor. For example, in this study, several participants identified the coffee smell as “chocolate” and lemongrass oil as “orange” or “phenyl flavor”. Hence the smell identification is considered as ‘normal’ when the subject either mentions the correct smell or any near perception. If the response is “no smell” or “undetectable” or “cannot identify” then it is considered as ‘anosmia’. The strip is made up of absorbent paper and if the response is “paper smell” then it is regarded as ‘anosmia’ for the odorized spots and ‘normal’ for the non-odorized spot (Position-2). Any smell response other than previously mentioned is considered as a distortion of smell or ‘parosmia’ for odorized spots and ‘phantosmia’ for the non-odorized spot.

### Statistical Analysis

Data were analyzed using IBM SPSS 20.0, Microsoft Excel, and visualized using R Studio *ggplot2* package. Descriptive statistics (numbers with percentages) were performed for categorical variables. Chi-squared tests were performed on categorical data as part of the secondary analysis. Multinomial regression analyses were performed on the categorical variables to assess the association between the COVID-19 test status and the sense of smell perception for all the six positions of the Anosmia checker. Logistic regression analysis was performed on the data collected from the fever clinic and suspected COVID cluster to find out the association between any osmia events to COVID-19 test positivity.

## Results

### Validation of ‘COVID-Anosmia Checker’ in COVID-19 positive patients and non-infected participants

Initially, we confirmed the performance of the test strip on 35 normal healthy individuals who were negative for COVID-19 by Antigen/Antibody testing **(Fig. 1)**. All the odorized spots (1, 3, 4, 5, and 6) were identified ‘exactly or similarly’ by 68-94% of the participants. The odorized spots were arranged in such a way that as the concentration of the odorized oil increased, the percentage of correct smell responses from the participants also increased (1.0µl coffee-71.4%, 2.0µl coffee- 88.6%, 5.0µl coffee- 68.6%, 5.0µl lemongrass oil- 94.3%, **Fig. 2A**). We observed a minor anosmia or parosmia response amongst healthy control participants who reported it as “non-identifiable” or “pleasant smell but cannot specify”. Parallelly, we conducted smell tests with the *COVID-Anosmia checker* on COVID-19 positive patients admitted at Saveetha Medical College Hospital, Chennai, India. The lowest coffee concentration (Position-3) was not recognized by 87.5% SARS-CoV-2 infected patients, which was correctly detected by 71.4% non-infected healthy individuals (**Fig. 2A**). It had been observed that higher the odor concentration less likely the COVID-19 participants were identified as anosmic (1.0µl coffee-Anosmia; OR=64.8, 95%CI= 18-233.4, p=0.000 and 5.0µl coffee – Anosmia; OR=25.6, 95%CI=8.3-78.1, p=0.000, **Table. 1**). This finding is consistent with the increasing events of hyposmia (5.3% and 10.5% hyposmia in Positions-5, 5.0µl Coffee oil and 6, 5.0µl Lemon grass oil respectively, **Fig. 2A**). Thus, it implies that other than complete loss of smell (anosmia) there are events of reduced smell or hyposmia that otherwise cannot be reliably detected by directly smelling household items. This is evident from the fact that only three COVID-19 positive patients in our cohort self-declared as having experienced the loss of smell.

**Figure 1:**
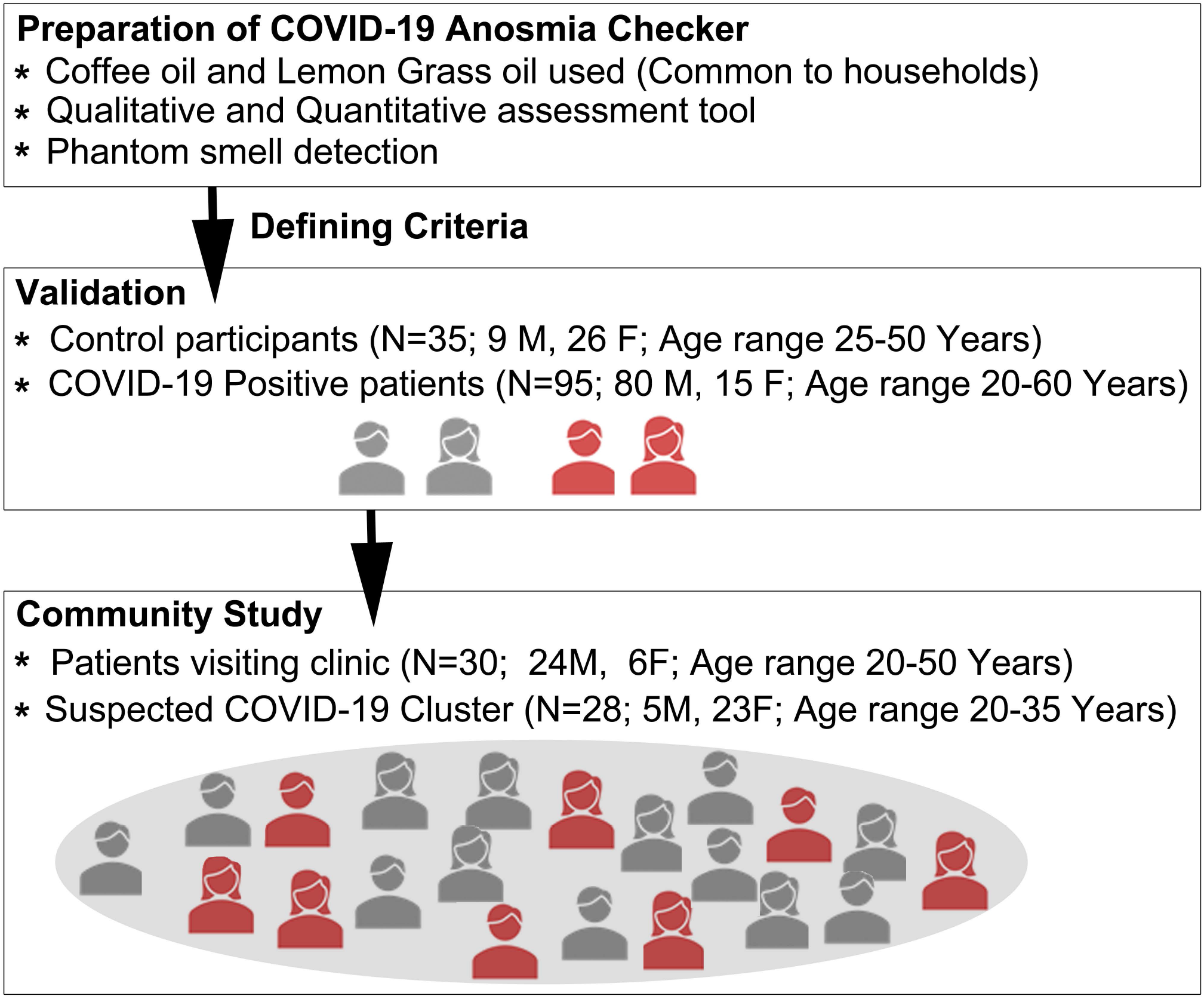
Flowchart illustrating the study design. It includes preparation of paper-based odorized strip (named as *COVID-Anosmia Checker*), through validation of the strip and finally community study for screening COVID-19 infection.

**Figure 2:**
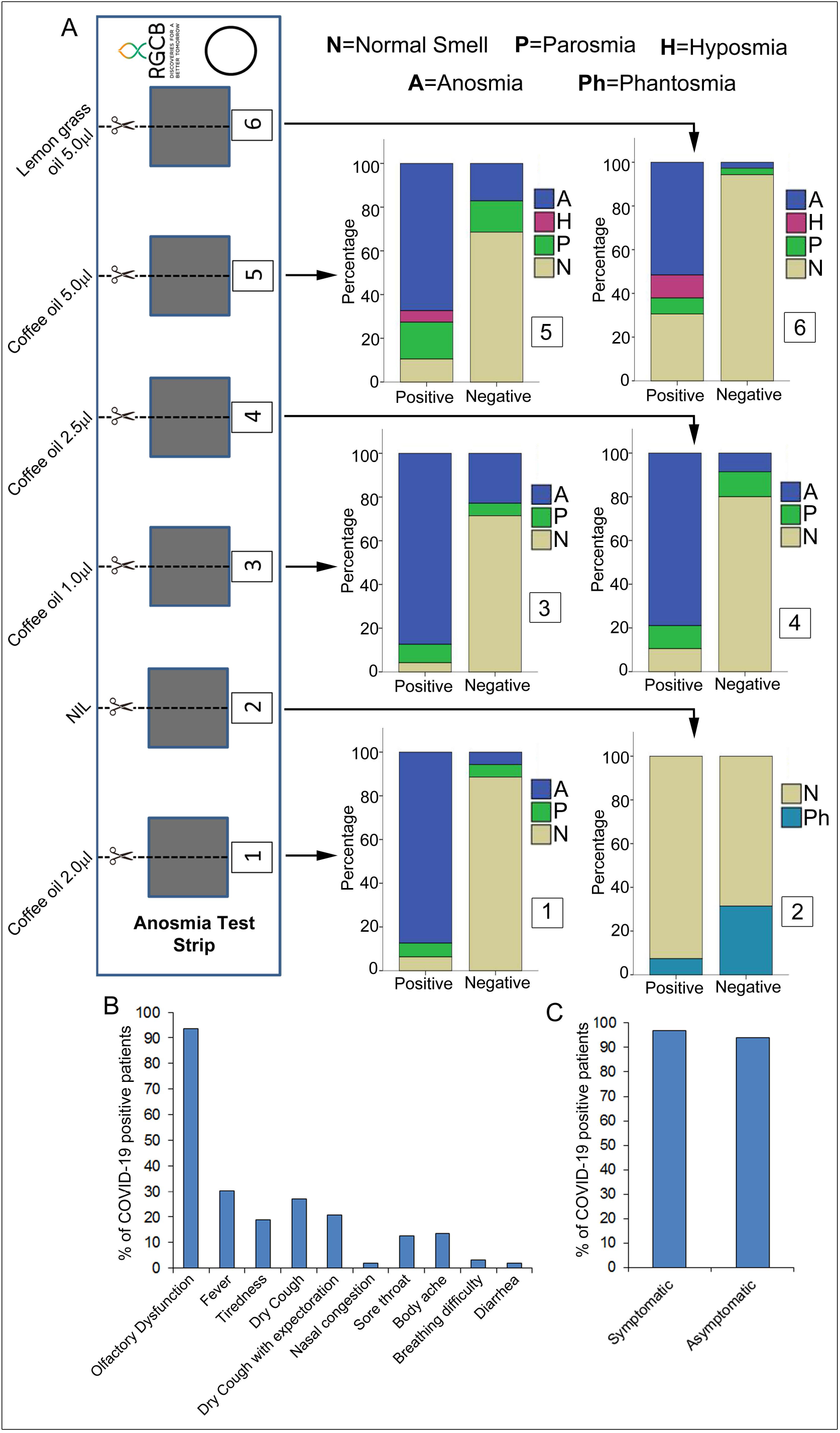
*COVID-Anosmia Checker* validation amongst healthy and patient subjects. (A) The smell perception response at each position of the strip is plotted as a stacked bar graph. (B) The bar graph shows the percentages of COVID-19 patients to have the symptoms mentioned. (C) The bar graph shows that 97% of the symptomatic and 94% of the asymptomatic COVID-19 positive patients had olfactory dysfunction including anosmia, hyposmia and parosmia.

**Table 1:** Result of Multinomial Logistic Regression analysis of being Anosmic/Parosmic/Hyposmic between COVID-19 positive vs COVID-19 negative participants.

| **Dependent Variable** | **Categories** | **Odds ratio** | **CI 95%** | **P value** |
| --- | --- | --- | --- | --- |
| **Position-1**<br>2.0µl Coffee oil | Normal | 1 |  |  |
|  | Anosmia | 214.4 | 41-1119 | 0 |
|  | Parosmia | 15.5 | 2.5-95.9 | 0.003 |
| **Position-2**<br>NIL | Phantosmia | 1 |  |  |
|  | No Smell | 5.7 | 2.0-16.4 | 0.001 |
| **Position-3**<br>1.0µl Coffee oil | Normal | 1 |  |  |
|  | Anosmia | 64.8 | 18-233.4 | 0 |
|  | Parosmia | 25 | 3.8-162.9 | 0.001 |
| **Position-4**<br>2.5µl Coffee oil | Normal | 1 |  |  |
|  | Anosmia | 70 | 17.9-273 | 0 |
|  | Parosmia | 7 | 1.7-27.4 | 0.005 |
| **Position-5**<br>5.0µl Coffee oil | Normal | 1 |  |  |
|  | Anosmia | 25.6 | 8.3-78.1 | 0 |
|  | Parosmia | 7.6 | 2.2-26.7 | 0.001 |
| **Position-6**<br>5.0µl Lemon grass oil | Normal | 1 |  |  |
|  | Anosmia | 55.7 | 7.2-429.5 | 0 |
|  | Parosmia | 7.9 | 0.9-68.6 | 0.05 |

CI, Confidence Interval

Phantosmia or phantom sensation of smell is relatively uncommon symptom seen in trivial illness affecting nasal epithelium as well as in many illnesses affecting central nervous system^23–26^. It is also reported to be present in healthy individuals ^27,28^. Here, we hypothesized that if a COVID-19 patient loses the smell sensation then the person might not show any phantom sensation. Quite surprisingly only 7.4% COVID-19 positive patients reported phantom smell at Position-2 (No odor) of the strip, which corresponded to those patients who had smell sensation normal to some extent. On the other hand, 31.4% of the control participants reported phantosmia as expected in general (**Fig. 2A**).

### Anosmia is the most prominent and salient symptom in both asymptomatic and symptomatic COVID-19 patients

Interpretation of the data showed that almost 95% of the COVID-19 positive patients showed some degree of olfactory dysfunction ranging from partial to complete loss of smell (**Fig. 2B**). The remaining 5% of the COVID-19 positive subjects showed normal smell perception indicating the possibility of the recovery from the disease. Symptom analysis showed that only 30% of COVID-19 positive patients reported fever, 27% dry cough, and 13% had body pain. However, comparatively lower proportion of the patients reported breathing difficulty (3%) and diarrhea (2%) (**Fig. 2B**). The loss of smell (95%) turned out to be the most common symptom amongst the positive patients which is consistent with the previous findings^14–17^

Moreover, we have taken into account both symptomatic as well as asymptomatic COVID-19 patients. Amongst the symptomatic patients, 97% of them showed smell perception dysfunction **(Fig. 2C)**. On the other hand, when the so-called ‘asymptomatic’ patients were tested, majority of them were found to have olfactory dysfunction (94%, **Fig. 2C**). In the current study the proportion of asymptomatic subject was only 34% of the total patient-participants. That is attributed to the fact that majority of patients with clinical symptoms only were admitted to the hospital (**Fig. S3B**). Practically, we expect to find an increased percentage of asymptomatic subjects compared to symptomatic patients in a community screening.

### Prediction of the COVID-19 test positivity using Anosmia checker tool in fever clinic and suspected COVID-19 cluster

Next, we wanted to verify whether the *COVID-Anosmia checker* could be used to predict the COVID-19 test positivity. We explored the association between the loss of smell and SARS-CoV-2 infection. We ran the anosmia test in a small set of population (N=58; age range 20-50) who were suspected to have COVID-19 or were primary contacts (**Fig. 1**). This group was a representative of a small community to check the efficiency of the *COVID-Anosmia checker* to initially screen and identify possible COVID-19 infection. Logistic regression analysis showed that a single altered response (anosmia/ parosmia/ hyposmia) in the smell test increased the likelihood of being tested positive for COVID-19 by 3.8 times (OR 3.8, 95%CI 1.5-9.4, p=0.004, **Table. 2**). Out of six spots in the *COVID-Anosmia checker* five of them are odorized and consistent altered smell response for all the five spots increases the probability to be tested positive for COVID-19.

**Table 2:** Logistics Regression Analysis exploring the probability of being tested positive for COVID-19 with ‘Sum of Osmia’ score

|  | **Odds ratio**<br>**(95% CI)**<br>**(unadjusted)** | **p-value** | **Odds ratio**<br>**(95% CI)**<br>**(adjusted)\*** | **p-value** |
| --- | --- | --- | --- | --- |
| Score of<br>Osmia | 3.80<br>(1.53-9.42) | 0.004 | 2.80<br>(1.17-6.66) | 0.020 |
| Constant | 0.002 | 0.005 | 0.001 | 0.008 |

CI, Confidence Interval;
\* For age and sex

In the community study, we categorized the participants (N=58) into three risk groups based on their smell response. Altered smell response for each of the five odorized spots was counted as score 1 into the ‘Sum of Osmia’ score. A ‘Sum of Osmia’ score was considered as ‘low risk group’ if the value is 0-2; ‘medium risk group’ for values 3-4 and ‘high risk group’ for value of 5. **(Fig. 3A)**. Out of 58 participants, we found that 26, 15, and 17 of them were at high-risk, medium-risk, and low-risk category respectively. 17 (65%) of the high-risk group, 4 (26%) of the medium-risk group were found to be COVID-19 positive through RT-PCR testing whereas none of the low-risk group participants turned positive indicating a 100% specificity.

**Figure 3:**
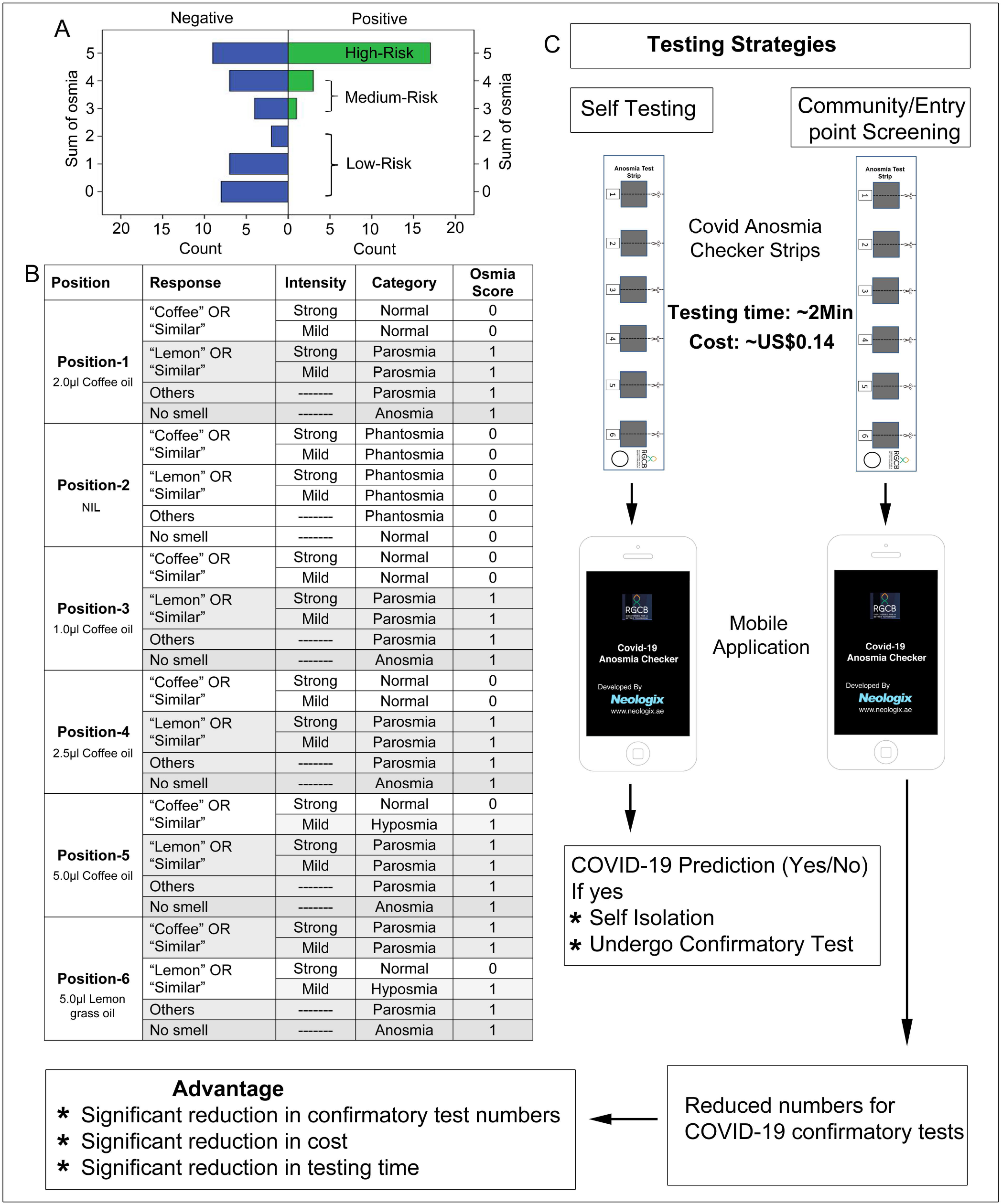
Mobile Application development for easy categorization of the user. Any deviation of response from the original odor is counted as score 1 into the sum of the Osmia score. Position-2 is excluded from the scoring system. (A) The maximum Sum of Osmia score is 5 for the five odorized positions. There are three risk categories based on the Sum of Osmia score used for the community study; Low risk group (0-2), medium risk group (3-4), high risk group (5). None of the low risk group category was tested positive for COVID-19 that indicates 100% specificity. (B) The table shows the logical scoring of the smell perception response used in the mobile application development. (C) The *COVID-Anosmia Checker* can be used as self-testing as well as community or entry point screening.

### COVID-Anosmia checker coupled with a mobile application for making unbiased decision on COVID-19 positivity

We have found that it would be difficult for a person who is not familiar with the test to make an unbiased interpretation of the Part-B scoring sheet. To overcome this issue, we developed a mobile application replacing the Part-B scoring sheet **(Fig. S2)**. The inputs for the application were statistically derived from the community screening as discussed earlier **(Fig. 3B, Table. 2)**. The person undergoing the test will have to enter the information as shown **(Fig. S2)**. Based on the detection of the odorized spots, the application will give the output with the ‘Sum of Osmia’ score as “Low-Risk” (0-2), “Medium-Risk” (3-4) or “High-Risk” (5). If a person is self-testing with the *COVID-Anosmia checker* and gets an output as “Medium Risk” or “High Risk” then there is a high probability for that person to be COVID-19 positive and should self-quarantine to reduce the spread **(Fig. 3C)**. Whereas, in community screening of suspected COVID-19 clusters or at airport/border entry points only persons with “Medium-Risk” or “High-Risk” need to be subjected to confirmatory COVID-19 tests thereby significantly reducing the test numbers and cost **(Fig. 3C)**.

## Discussion

To counter the shrinking economy, countries throughout the world have no other option but to lift the lockdown in a graded manner. Various countries including India have started lifting the lockdown once the dynamics of the virus and its case fatality rate were understood^29^. This poses a threat to the health sectors as the second wave of infection is being reported in several parts of the world including the UK, France, Spain, etc. (https://coronavirus.jhu.edu/data/new-cases). These circumstances are making it even more critical to self-test and self-isolate than ever before to limit the spread of SARS-CoV-2 virus. So far there is no available test kit to self-test reliably for SARS-CoV-2 infection. As loss of smell is a recently recognized symptom of COVID-19 ^21^, developing a smell test kit was the need of the hour. We noted only one relevant finding^30^ that developed olfactory action meter which requires technical sophistication and expert staff to operate and interpret. None of the reports mentioned about developing any tool to detect loss of smell that is cost-effective and can be readily used by any individual without necessary expertise. Here, we developed a low-cost olfactory assessment tool and validated it amongst the COVID-19 positive and negative participants.

We found that quantitative assessment of smell perception is equally crucial as reduced smell or hyposmia may be underestimated while self-testing with available household items. It is important to note that when participants reported reduced smell or hyposmia, in each case their COVID-19 test report turned out to be positive although such number is less in our cohort study. Finally, we have shown that the *COVID-Anosmia checker* can be used to predict the SARS-CoV-2 infection. Currently, we rely heavily on thermal screening alone to look for symptomatic individuals that entail around 14% of the COVID-19 infected individuals and are significantly missing the rest **(Fig. S3A)**. Based on our findings we suggest a radical change in COVID-19 screening protocol in public places such as border entry points, schools, restaurants, and airports. It would be ideal to screen out large communities with olfactory dysfunction using the *COVID-Anosmia checker* along with thermal screening and further, the smaller high or medium-risk group can be subjected to confirmatory COVID-19 testing platforms **(Fig. 3C)**. This protocol will be extremely useful for low and middle-income countries since it can significantly reduce the proportion of patients that needs to be subjected to costly RT-PCR based confirmation. A model for community screening is depicted in **Fig. 3C**. Although detection through RT-PCR remains the gold standard of COVID-19 detection, *COVID-Anosmia checker* can be a rapid alternative for self-testing at home which is inexpensive and less time-consuming. An individual can repeatedly self-test at home to check for the suspected onset of COVID-19. The primary screening using the *COVID-Anosmia checker* is swift and takes only ∼2 minutes to complete the test and costs ∼US$ 0.14 for manufacturing each test strip. Our current version of the odorized test strip has a shelf life of ∼1month. Trapping the same concentrations of odors in blister packs, similar to those used for packing tablets can further increase the shelf life. These blister strips will also help in easy pop-opening of the blisters with the fingers.

Further, coupling the *COVID-Anosmia checker* with the mobile application helped in easy unbiased categorization of the risk groups. Currently, the application can be downloaded onto any android smartphone and used as standalone software. The current android version has the potential to be further integrated with any health systems software to generate a database to track and quarantine the high-risk group.

Based on our above findings we stress the usefulness of the *COVID-Anosmia checker* as a low-cost, fast and accurate quantitative primary mass-screening tool, combined with confirmatory platforms or for self-checking and isolation. It can also be effectively used in difficult to reach geographical locations. This approach could significantly reduce cost, time and enhance detection and tracing of symptomatic and asymptomatic COVID-19 patients thereby immensely helping in management and control of COVID-19 spread.

## Data Availability

We agree to make available materials, data and associated protocols used in this study upon request.

## Authors’ Contributions

Conceived the study **BB, JJ**; Developed the study protocol **BB, PAR, SP, JI, BSN, JJ**; Anosmia checker tool generation **BB, PAR, SP, JI, BSN, SG, PT, JJ**; Data collection **BB, SP, DSK, PRK, GH, MJ, PKS, LDVN, RV, RK, BK, IJ, RN**; Data analysis and curation **BB, APR, JJ**; Mobile application development **GKA**; Writing original draft **BB, APR, JJ**; Research Coordination **JJ, MRP**; Funding acquisition **JJ**

## Acknowledgments

This work was supported by Intramural grants to J.J. from Rajiv Gandhi Centre for Biotechnology (RGCB) and external funding from the Department of Science and Technology (SERB), Government of India (CVD/2020/001011). B.B (UGC-332486), P.A.R (CSIR-09/716(0156)/2015-EMR-I) and S.P (CSIR-09/716(0161)/2015-EMR-I) were supported by research fellowship from Government of India. We thank Spiceor Bionutralites Pvt. Ltd., Cochin, Kerala for providing the Coffee oil. We thank Sreedevi S (RGCB) and Vision Graphics, Trivandrum for manufacturing the test strips. We thank Dr. Sudhesh Kumar Kattumannil (Indian Statistical Institute, Chennai) for suggestions over statistics and figures. Thanks are also due to Dr. Ani V Das (RGCB) for critically evaluating the manuscript.

## Figure Legends

**Figure S1: COVID-19 Anosmia Checker Details**. (A) Anosmia paper strip shows how absorbent paper pieces are glued to the non-absorbent paper in a row numbered as 1 to 6. Each absorbent paper is spotted onto the middle of the dotted line with different volumes of odorant oils owing to create a gradient of odors. The second position is left blank. (B) The form associated with the paper strip contains the details to be filled by the subjects.

**Figure S2: *COVID-Anosmia Checker* mobile application**. The screenshot of the mobile application developed by Neologix software solutions.

**Figure S3: COVID-19 symptoms proportions**. (A) The pie diagram represents the proportions (in scale of 100) of the classic symptoms of COVID-19 along with olfactory dysfunction. (B) The pie chart shows the proportions of the ‘symptomatic’ and ‘asymptomatic’ subjects included in the study.

