## Supplementary material for "COVID-Anosmia Checker: A rapid and low-cost alternative tool for mass screening of COVID-19": Fig. S

Supplementary Fig-1

A

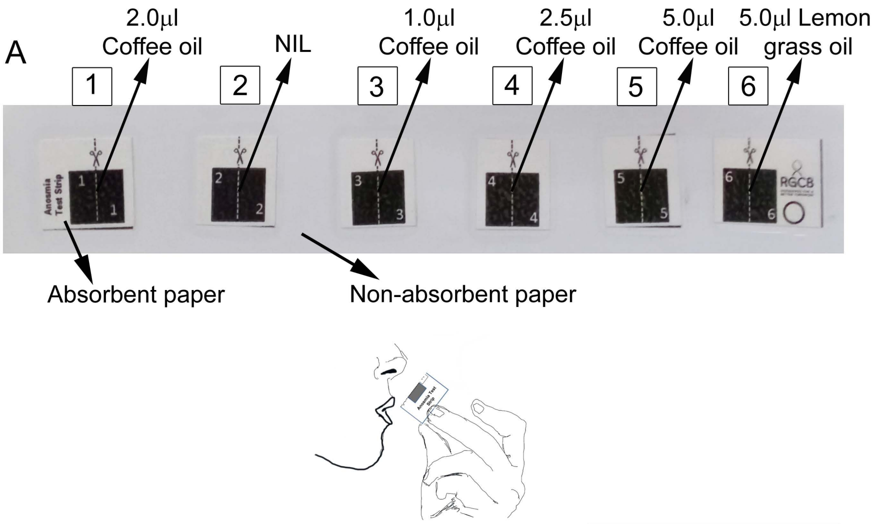


B

RGCB
DISCOVERIES FOR A BETTER TOMORROW

**ID No.** ☐

### COVID-Anosmia Checker

**Instructions for carrying out COVID-Anosmia Checker Test**

1. Please fill Item No. ❶ & ❸ completely.
2. Cut the anosmia strip along the cut mark in **Box 1**. Smell for odor along the cut end. Tick YES if you smell an odor and NO if you don't smell an odor in Item No. ❷. Please also identify the odor that you smell. Repeat the same for **Box No. 2-6**.

❶

**Name:** ____________________

**Address:** ____________________

____________________

**Phone No:** ____________________

**Did you travel outside India within the past 14 days? YES** ☐ **NO** ☐ **Age** ☐

**If yes then please provide the following details.**

**Date of Travel:** ____________________

**Flight No:** ____________________

**Passport No:** ____________________

**Gender M/F** ☐

❷ Please mark your answer with a ☑

| | Yes | No | Identify the odor? If No the write NIL |
|---|---|---|---|
| ① | ☐ | ☐ | ☐ |
| ② | ☐ | ☐ | ☐ |
| ③ | ☐ | ☐ | ☐ |
| ④ | ☐ | ☐ | ☐ |
| ⑤ | ☐ | ☐ | ☐ |
| ⑥ | ☐ | ☐ | ☐ |

❸ **Do you have any of the following symptoms?**

| | | Date of onset |
|---|---|---|
| a) Fever | ☐ | ☐ |
| b) Tiredness | ☐ | ☐ |
| c) Dry Cough | ☐ | ☐ |
| d) Cough with expectoration | ☐ | ☐ |
| e) Sore throat | ☐ | ☐ |
| f) Body aches and pains | ☐ | ☐ |
| g) Nasal congestion | ☐ | ☐ |
| h) Runny nose | ☐ | ☐ |
| i) Diarrhea | ☐ | ☐ |
| j) Breathing difficulty | ☐ | ☐ |
| k) Loss of smell | ☐ | ☐ |
| l) Loss of taste | ☐ | ☐ |
| m) None of the above | ☐ | |

**To be filled by appropriate testing authority**

**Date of Test:**

**Name:**

**Signature:**

**To be filled by RGCB**

**Anosmia Checker Result** **Passed:** ☐ **Failed:** ☐

**RT-PCR Result**

**Positive:** ☐

**Negative:** ☐

**Date of testing** ☐

**Antibody Test Result**

**Positive:** ☐

**Negative:** ☐

**Date of testing** ☐

○

**Name:**

**Signature:**

Supplementary Fig-2

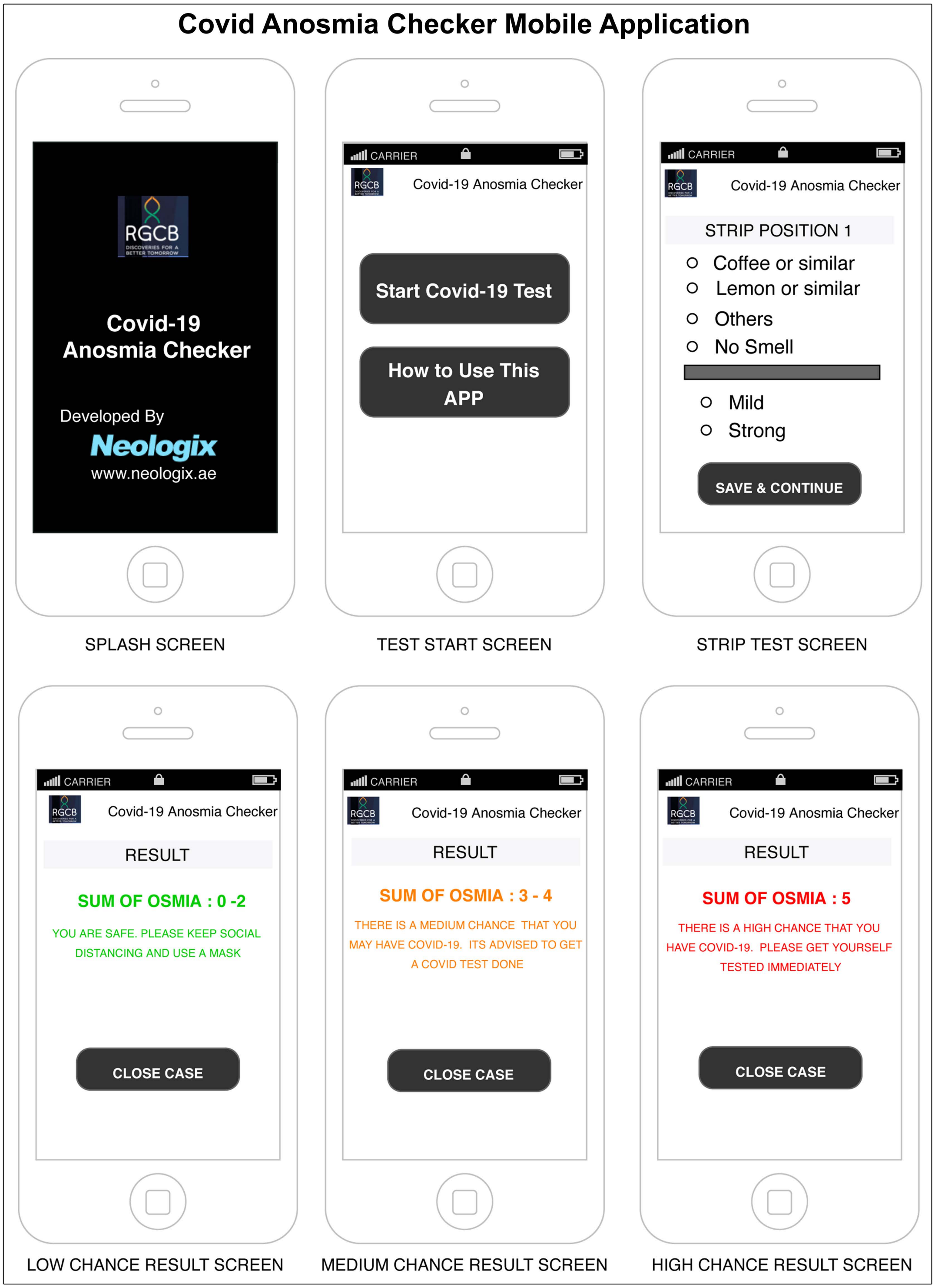

Supplementary Fig-3

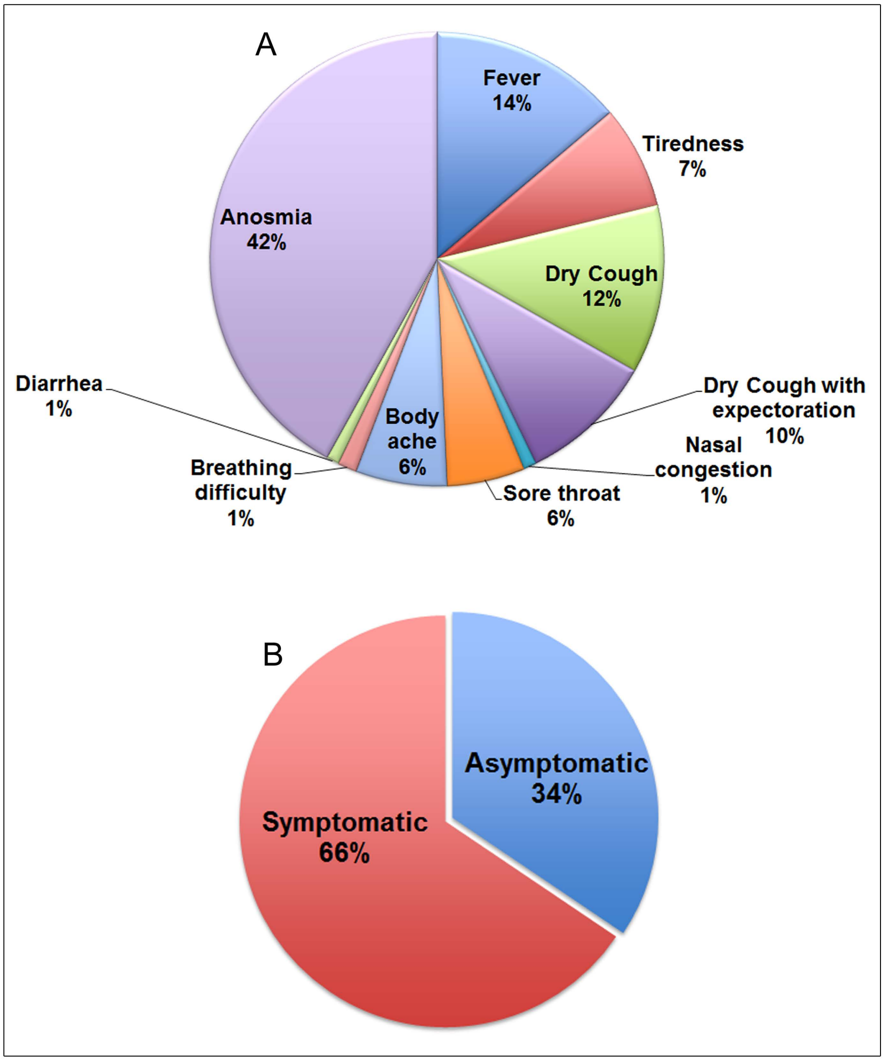
